# Pathology testing for patients with low back pain in Australian emergency departments

**DOI:** 10.1101/2025.09.07.25335026

**Authors:** Claudia Côté-Picard, Qiuzhe Chen, Hugo Massé-Alarie, Peter Youssef, Michael Spies, Chris G Maher, Gustavo C Machado

**Author notes:** **Corresponding author:** Professor Gustavo Machado, Sydney School of Public Health, Faculty of Medicine and Health, The University of Sydney, Sydney, New South Wales, Australia.

## Abstract

**Background:** Imaging in low back pain management is well documented, but little is known about pathology testing in this population. We aimed to describe the profile of patients with low back pain who received pathology testing in emergency departments, and to describe the ordered tests.

**Methods:** A retrospective study of electronic medical records from three emergency departments in Sydney, New South Wales, Australia, from January 2016 to October 2021, was undertaken. We included patients diagnosed with a lumbar spine condition at discharge from the emergency department and extracted their demographic and episode of care characteristics. We performed logistic regressions adjusted for age, sex and triage category to identify the factors associated with pathology testing.

**Results:** Pathology tests were ordered in 23.8% of 15 300 episodes of care. Patients who were admitted (adjusted odds ratio (aOR) 60.54, 95% CI 52.5, 69.81), stayed longer in the emergency department (aOR 3.76, 95% CI 3.45, 4.10), had a serious pathology (aOR 2.74, 95% CI 2.30, 3.27), or arrived by ambulance (aOR 2.64, 95% CI 2.42, 2.87), were more likely to receive a pathology test than their counterparts. Full blood count, electrolytes, urea, creatinine (EUC), and liver function tests were the most ordered tests.

**Conclusions:** The characteristics associated with pathology testing were similar to those reported for lumbar imaging in patients with low back pain in previous studies. Our results suggest that guideline recommendations for pathology testing were partly followed, but an investigation into the appropriateness of pathology testing is needed to confirm this hypothesis.

## Introduction

Low back pain (LBP) is a common reason for emergency department (ED) visits. A meta-analysis of 21 studies from 12 countries estimated that the prevalence of LBP in the ED was 4.4% of cases (1). About 45% of ED presentations for LBP have diagnoses of a lumbar spine condition, and of those, most are non-urgent (2). The majority (85%) are non-specific, 10% are radicular syndromes, and only 5% are serious spinal pathologies (e.g. epidural abscess) (2).

Diagnostic work-up including pathology testing and imaging should be reserved for LBP patients for whom a serious pathology is suspected or when a surgical intervention is considered (3). Most guidelines for LBP provide advice on imaging, but guidance on pathology testing is less commonly provided (4). In Australia, the Colleges for Emergency Medicine and Pathology have recently released a joint guideline for pathology testing in the ED (5). The tests recommended for admitted patients with non-traumatic LBP are urea, creatinine, electrolytes and glucose, full blood count, and dipstick urinalysis. The Emergency Care Institute, Australia, recommends that pathology tests be ordered when a specific lumbar spine pathology or a non-spinal cause is suspected in acute LBP (6).

Lumbar imaging use for patients with LBP has been well documented. A meta-analysis of 4 studies including 16 552 LBP episodes of care estimated a 36% rate of imaging in ED (7). An Australian study of 649 patients presenting to ED with LBP reported imaging overuse and underuse prevalences of ∼9% and ∼4%, respectively (8), while another with 1 459 patients reported a 4.8% overuse rate (9). In contrast, only a few studies described pathology testing use in LBP management. The ordering rate for pathology testing in ED care of LBP was ∼20% in studies from Canada and the US (10, 11), 44% in patients 65 years and older in Australian EDs (12), and 62.2% in a private Australian ED (13). There remains a lack of evidence regarding the profile of patients with LBP who are ordered pathology tests in EDs, and the characteristics of the ordered tests. The aims of this study were to describe the profile of patients with LBP who were ordered pathology tests in three Australian EDs, the types of pathology tests ordered, and to investigate the factors associated with the ordering of a test.

## Methods

### Study design, setting and population

This is a retrospective analysis of ED electronic medical record data of patients aged 18 years and older who presented to one of three major public hospitals within the Sydney Local Health District, New South Wales, Australia (Royal Prince Alfred Hospital, Concord Repatriation General Hospital and Canterbury Hospital) from January 2016 to October 2021 and who received a SNOMED-CT diagnosis code related to a lumbar spine condition at discharge from ED (14).

The Sydney Local Health District (Royal Prince Alfred Hospital zone) Ethics Committee granted approval for this study (protocol no. X17-0419). Patients who presented with LBP but received a non-lumbar spine diagnosis at discharge from ED were not included. We followed the Reporting of studies Conducted using Observational Routinely-collected Data (RECORD) guideline (15, 16).

### Study procedures

Electronic medical record data were obtained by the Sydney Local Health District Targeted Activity and Reporting System (STARS), that routinely collects data from patients with LBP in the three EDs (16).

#### Demographic and episode of care characteristics

We extracted patient demographic and episode of care characteristics. Demographic characteristics were age, sex, postcode, country of birth (Australia or overseas), preferred language (English or other), and whether the patient required an interpreter. Episode of care characteristics were the mode of arrival in ED (ambulance or other means), presentation hour and day, ED triage category, diagnosis at discharge from ED, whether there was a hospitalisation, whether there was a re-presentation to the same ED within 28 days, and ED length of stay.

Socioeconomic status was derived from the patient’s postcode using the Australian Bureau of Statistics’ Index of Relative Socioeconomic Advantage and Disadvantage (IRSAD) for Areas 2016 (17). IRSAD were extracted as deciles and dichotomised to disadvantaged (1–5) or advantaged (6–10). ED visits from Monday to Friday between 8:00 and 17:00 were considered as being during working hours. ED clinicians triaged the patients using the Australasian Triage Scale (18), that ranges from 1 to 5, and we dichotomised it to urgent (1–3) and less urgent cases (4–5). Lumbar spine diagnoses were grouped into two categories: non-serious LBP, including non-specific and radicular LBP, and LBP due to a serious spinal pathology (serious LBP). ED length of stay was the time in hours from ED arrival to discharge or hospital admission. When the length of stay was greater than 4 hours, it was categorised as extended. For 1926 LBP episodes, an admission was recorded but data was missing for hospital ward and date and time of admission. We were able to verify one third of these presentations with another data set, and those verified were solely admitted to ED short stay units, which are designed for stays shorter than 24 hours, and so we considered these 1926 episodes as non-admitted.

#### Pathology tests

We extracted all pathology tests that were ordered at least once, and two rheumatologists (MS and PY) sorted the ordered tests into nine categories. We excluded variables that were groups of tests as the individual tests included in these groups were also recorded individually. We excluded variables that were recorded as pathology tests and that represented a non-test process that was not ordered by the clinical team (e.g. “_POC RPA Billing”, “Haem Comment”, “Coagulation Comment”). The classification of pathology tests into categories and the excluded tests are reported in the Supplementary Table S1.

### Outcome measures

Our outcomes were: (i) the proportion of LBP episodes where at least one pathology test was ordered, by demographic and episode of care strata; (ii) the associations between demographic/ episode of care characteristics and pathology test orders; (iii) the proportion of LBP episodes who were ordered pathology tests in different categories: biochemistry, endocrinology, immunology, haematology, coagulation, blood gas, microbiology, oncology, and drug monitoring – toxicology, and by LBP diagnosis at discharge from ED (serious vs non-serious) and admission (yes/ no) strata; (iv) the 10 most frequently ordered pathology tests, by LBP diagnosis and admission strata, and (v) the distribution of the number of ordered pathology tests by LBP diagnosis and admission strata.

### Data analysis

We calculated the number of LBP episodes where a pathology test was ordered, with results stratified by demographic/ clinical characteristics. Categorical variables are presented as frequency (%) and continuous variables as mean (SD). Multiple presentations for LBP by a same patient over the six-year study period were analysed independently as different presentations.

Multivariable logistic regressions were performed to assess the association between demographic/ clinical characteristics and the order of at least one pathology test, adjusting for age, sex and triage category. The covariates were chosen based on the team expertise and on the available literature regarding the factors associated with pathology testing and diagnostic work-up in emergency care of LBP (2, 12). We reported the results of these models as adjusted odds ratio (aOR) and 95% confidence interval (CI). The effect sizes were interpreted as small (1.5≤ aOR <2.0 or 0.5< aOR ≤0.67), medium (2≤ aOR <3 or 0.33< aOR ≤0.5) or large (aOR ≥3 or ≤0.33) (19). An aOR< 1.5 or >0.67 was considered as non-important. The analyses were conducted in Stata/BE 18.0.

## Results

### Patient characteristics and pathology test orders

This study included 15 300 LBP episodes of 11 521 unique patients who presented to one of three EDs and who received an ED discharge diagnosis related to the lumbar spine from January 2016 to October 2021. The characteristics of the patients and their ED episodes are presented in Table 1. The sample included 7 775 (50.8%) females, and the mean age was 52.9 years ± SD 20.1. Most diagnoses at discharge from ED were non-serious LBP, with 14 642 episodes (95.7%).

**Table 1.** Demographic and episode of care characteristics.

|  | Overall | Any laboratory test | No laboratory test |
| --- | --- | --- | --- |
| **Demographic characteristics** |  |  |  |
| Number of episodes, n (%) | 15 300 (100) | 3 634 (23.8) | 11 666 (76.3) |
| Age (years), mean [SD] | 52.9 [20.1] | 65.6 [19.3] | 48.9 [18.7] |
| <50, n (%) | 7 236 (47.3) | 804 (11.1) | 6 432 (88.9) |
| 50 to 65, n (%) | 3 434 (22.4) | 762 (22.2) | 2 672 (77.8) |
| >65, n (%) | 4 630 (30.3) | 2 068 (44.7) | 2 562 (55.3) |
| **Sex\*** |  |  |  |
| Female, n (%) | 7 775 (50.8) | 2 084 (26.8) | 5 691 (73.2) |
| Male, n (%) | 7 523 (49.2) | 1 549 (20.6) | 5 974 (79.4) |
| **Socioeconomic status – IRSAD<sup>†</sup>** |  |  |  |
| Disadvantaged (1-5), n (%) | 7 772 (53.7) | 1 880 (24.2) | 5 892 (75.8) |
| Advantaged (6-10), n (%) | 6 699 (46.3) | 1 685 (25.2) | 5 014 (74.9) |
| **Country of birth<sup>‡</sup>** |  |  |  |
| Australia, n (%) | 7 158 (47.1) | 1 618 (22.6) | 5 540 (77.4) |
| Overseas, n (%) | 8 043 (52.9) | 2 003 (24.9) | 6 040 (75.1) |
| **Preferred language<sup>§</sup>** |  |  |  |
| English, n (%) | 11 392 (74.5) | 2 491 (21.9) | 8 901 (78.1) |
| Non-English language, n (%) | 3 906 (25.5) | 1 143 (29.3) | 2 763 (70.7) |
| **Interpreter required** |  |  |  |
| Yes, n (%) | 1 320 (8.6) | 502 (38.0) | 818 (62.0) |
| No, n (%) | 13 980 (91.4) | 3 132 (22.4) | 10 848 (77.6) |
| **Episode of care characteristics** |  |  |  |
| **Mode of arrival** |  |  |  |
| Ambulance, n (%) | 5 064 (33.1) | 2 029 (40.1) | 3 035 (59.9) |
| Other means, n (%) | 10 232 (66.9) | 1 605 (15.7) | 8 627 (84.3) |
| **Time of arrival** |  |  |  |
| Working hours presentation<sup>¶</sup>, n (%) | 6 998 (45.7) | 1 879 (26.9) | 5 119 (73.2) |
| Outside of working hours, n (%) | 8 302 (54.3) | 1 755 (21.1) | 6 547 (78.9) |
| Triage category<sup>#</sup> |  |  |  |
| Urgent (1-3), n (%) | 6 420 (42.0) | 2 185 (34.0) | 4 235 (66.0) |
| Less urgent (4-5), n (%) | 8 879 (58.0) | 1 449 (16.3) | 7 430 (83.7) |
| LBP diagnosis at discharge from ED |  |  |  |
| Non-serious LBP**, n (%) | 14 642 (95.7) | 3 336 (22.8) | 11 306 (77.2) |
| Serious LBP, n (%) | 658 (4.3) | 298 (45.3) | 360 (54.7) |
| Admitted to hospital |  |  |  |
| Yes, n (%) | 2 647 (17.3) | 2 372 (89.6) | 275 (10.4) |
| No, n (%) | 12 653 (82.7) | 1 262 (10.0) | 11 391 (90.0) |
| Re-presentations within 28 days, n (%) |  |  |  |
| Yes, n (%) | 1 112 (7.3) | 265 (23.8) | 847 (76.2) |
| No, n (%) | 14 188 (92.7) | 3 369 (23.8) | 10 819 (76.3) |
| Extended length of stay (>4 hours), n (%) |  |  |  |
| Yes, n (%) | 4 221 (27.6) | 1 969 (46.7) | 2 252 (53.4) |
| No, n (%) | 11 079 (72.4) | 1 665 (15.0) | 9 414 (85.0) |
| Length of stay in ED (hours), mean [SD] | 4.2 [3.0] | 6.1 [4.1] | 3.6 [2.2] |
| * Sex was indeterminate for 2 low back pain episodes.<br>† IRSAD was missing for 829 episodes due to missing postcode, living overseas, no fixed abode or unspecified address.<br>‡ Country of birth was missing for 99 episodes.<br>§ Preferred language was missing for 2 episodes.<br>\|\| Mode of arrival information was missing for 4 episodes.<br>¶ Working hours are defined as being from 8:00 to 17:00 from Monday to Friday.<br># Triage category was missing for 1 episode.<br>** Non-serious LBP includes episodes with diagnosis at discharge of non-specific LBP and radicular LBP. |  |  |  |

At least one pathology test was ordered for 3 634 of 15 300 LBP episodes (23.8%) (Table 1). The number of individual types of tests recorded was 261, and 38 were excluded (9 non-tests and 29 groups of tests), leaving 223 different tests included in the analyses. Patients who received a pathology test were older (65.6 years ± SD 19.3) and stayed longer in ED (6.1 hours ± SD 4.1) than those who did not. Females were more likely to receive a pathology test than males (26.8% vs 20.6%), as were patients who had a preferred language other than English (29.3% vs 21.9%) and those who required an interpreter (38.0% vs 22.4%). Likewise, those who arrived by ambulance (40.1% vs 15.7%) and those arriving during working hours (26.9% vs 21.1%) were more likely to receive a pathology test. Those who were triaged as urgent (34.0% vs 16.3%) and those who received a serious LBP diagnosis at discharge from ED (45.3% vs 22.8%) were more likely to receive a pathology test. Those who were admitted to hospital were almost always ordered pathology testing (89.7%).

After adjusting for age, sex and triage category, the odds of pathology tests being ordered were higher for the episodes leading to hospital admission (aOR 60.54, 95% CI 52.5, 69.81), or with an extended ED length of stay (aOR 3.76, 95% CI 3.45, 4.10), with a large effect size (Table 2). Those who arrived to ED by ambulance (aOR 2.64, 95% CI 2.42, 2.87) and those who had a serious LBP diagnosis at discharge from ED (aOR 2.74, 95% CI 2.30, 3.27) also had higher odds of receiving pathology testing with a medium effect size. The odds of pathology tests being ordered were higher for those who were born in Australia (aOR 1.19, 95% CI 1.10, 1.30), required an interpreter (aOR 1.22, 95% CI 1.07, 1.39), or presented during working hours (aOR 1.21, 95% CI 1.12, 1.31) in a statistically significant manner, but with a non-important effect size (aOR <1.5).

**Table 2.**
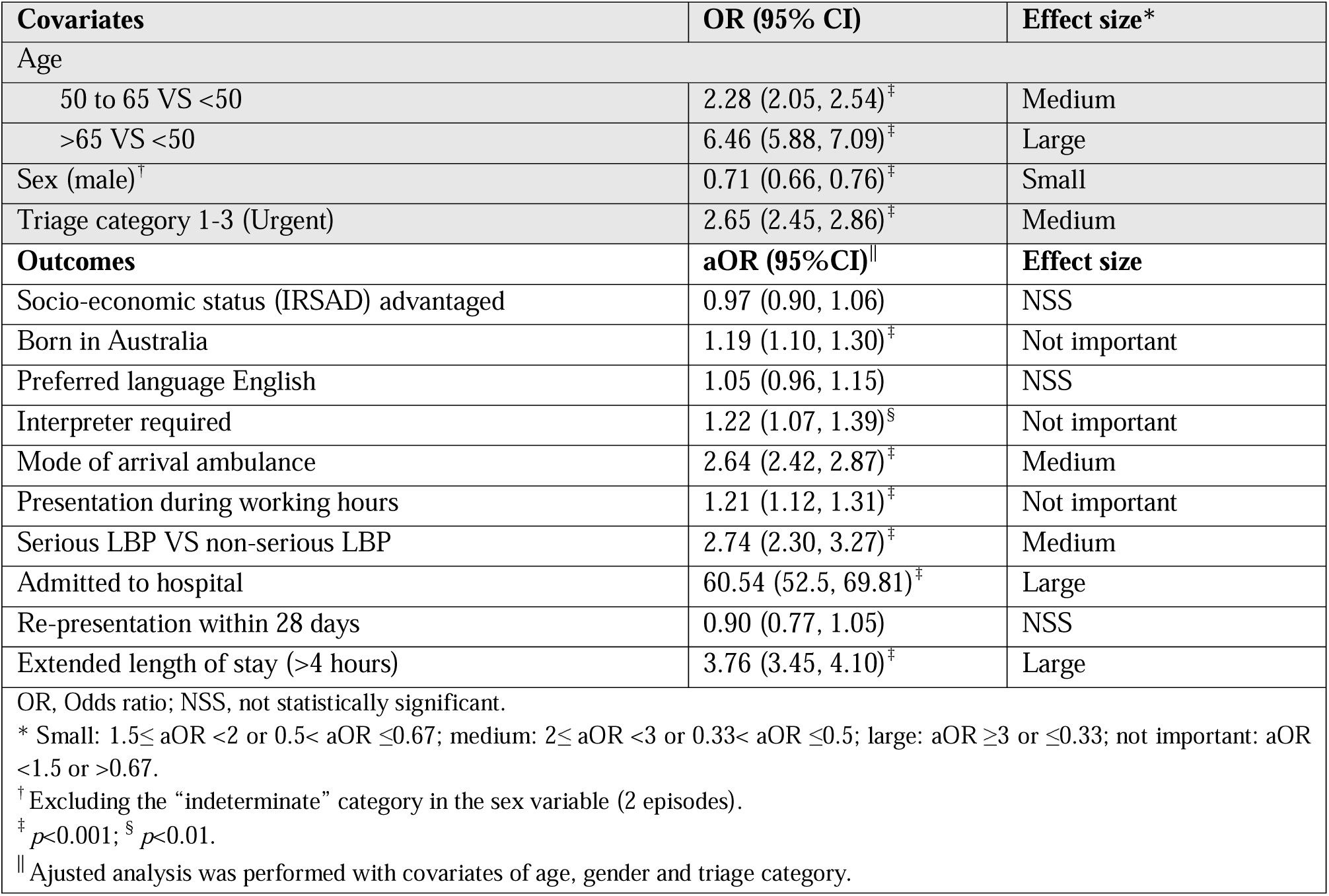
Association between laboratory test orders and demographic and clinical characteristics.

### Pathology test characteristics

Biochemistry, haematology, immunology, coagulation and blood gas were the five most ordered test categories (Table 3). The five most ordered pathology tests were full blood count, electrolytes, urea, creatinine, liver function tests, C-Reactive Protein, and calcium, magnesium, phosphate (Table 4). Fig. 1 shows the number of laboratory tests ordered for admitted and non-admitted episodes. For admitted episodes the mode was 6 pathology tests (with a 12.5% order rate) and the mode for non-admitted episodes was 5 tests (with a 1.7% order rate). Fig. 2 shows the number of laboratory tests ordered for serious and non-serious LBP episodes. The mode was 6 for both serious and non-serious LBP episodes (with a 5.5% order rate for serious LBP and 3.4% for non-serious LBP episodes).

**Fig. 1.**
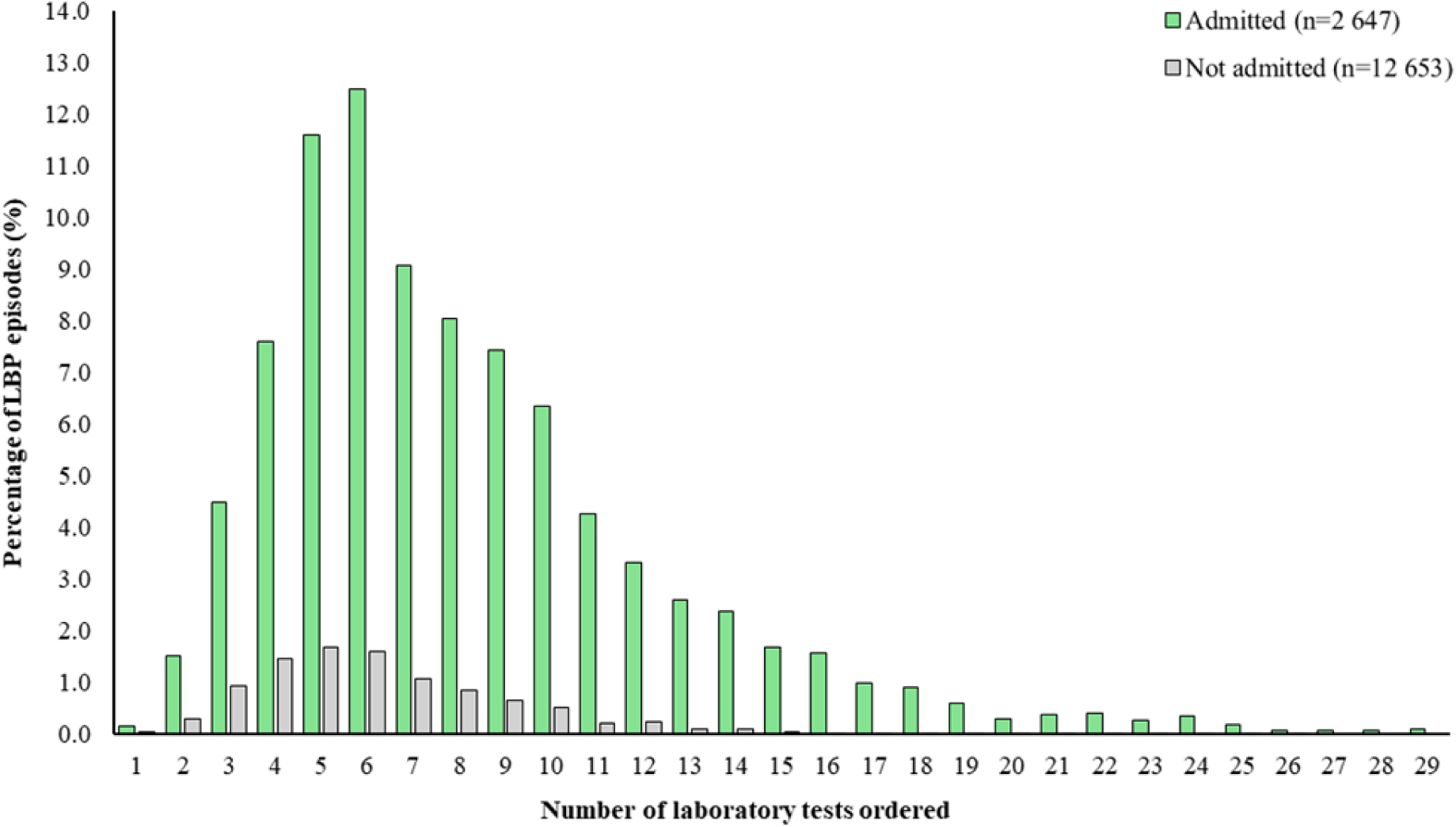
Percentage of LBP episodes per number of ordered tests for admitted (on a total of n=2 647) and non-admitted LBP episodes (on a total of n=12 653). The data for admitted LBP episodes who received 35, 36, 38, 40, 42, 44 and 100 laboratory tests were excluded as there was only one LBP episode who received each of these numbers of laboratory tests The data for 0 laboratory test ordered is not illustrated.

**Fig. 2.**
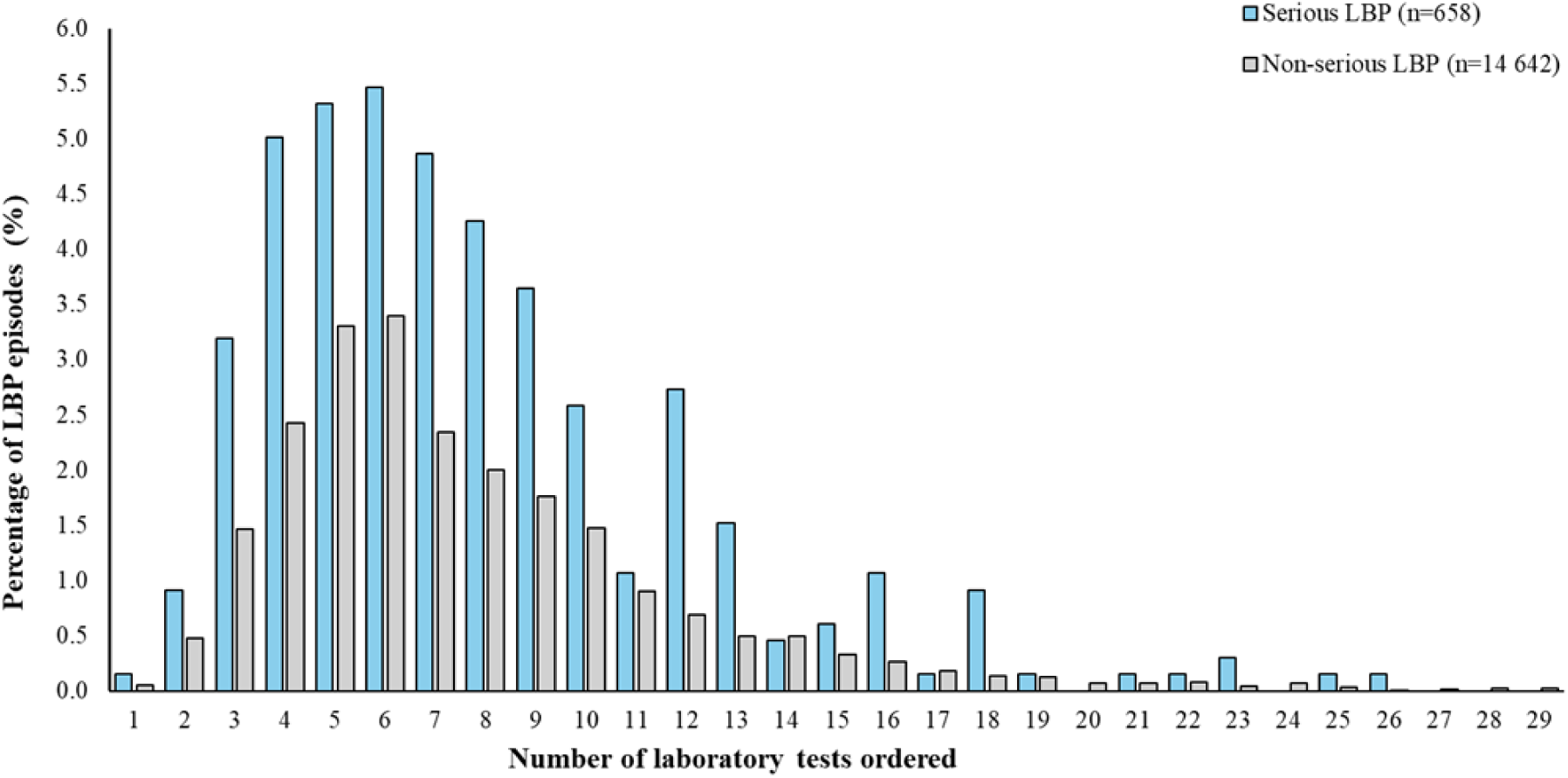
Percentage of LBP episodes per number of ordered tests for serious (on a total of n=658) and non-serious (on a total of n=14 642) LBP episodes. The data for non-serious LBP who received 35, 36, 40, 42 and 100 laboratory tests, and for serious LBP who received 38 and 44 laboratory tests were excluded as there was only one episode who received each of these numbers of laboratory tests. The data for 0 laboratory test ordered is not illustrated.

**Table 3.** Laboratory test categories ordered according to triage and low back pain diagnosis.

|  |  | LBP diagnosis at discharge from ED |  | Admitted to hospital |  |
| --- | --- | --- | --- | --- | --- |
|  |  | Non-serious LBP, n (%) | Serious LBP, n (%) | Yes, n (%) | No, n (%) |
|  | N | 14 642 | 658 | 2 647 | 12 653 |
| **Category (n of tests)** | **n** |  |  |  |  |
| 1. Biochemistry (58) | 3 619 | 3 321 (22.7) | 298 (45.3) | 2 365 (89.4) | 1 254 (9.9) |
| 2. Haematology (24) | 3 614 | 3 318 (22.7) | 296 (45.0) | 2 363 (89.3) | 1 251 (9.9) |
| 3. Immunology (33) | 2 579 | 2 401 (16.4) | 178 (27.1) | 1 744 (65.9) | 835 (6.6) |
| 4. Coagulation (10) | 993 | 886 (6.1) | 107 (16.3) | 744 (28.1) | 249 (2.0) |
| 5. Blood Gas (8) | 742 | 669 (4.6) | 73 (11.1) | 487 (18.4) | 255 (2.0) |
| 6. Endocrinology (21) | 586 | 523 (3.6) | 63 (9.6) | 423 (16.0) | 163 (1.3) |
| 7. Microbiology (48) | 231 | 199 (1.4) | 32 (4.9) | 191 (7.2) | 40 (0.3) |
| 8. Toxicology (15) | 135 | 110 (0.8) | 25 (3.8) | 94 (3.6) | 41 (0.3) |
| 9. Oncology (6) | 9 | 8 (0.1) | 1 (0.2) | 9 (0.3) | 0 (0) |
| n, number of episodes who received at least one test in the given category; N, number of episodes in the given admission to hospital or LBP diagnosis category.<br>% is the percentage of n/ N. |  |  |  |  |  |

**Table 4.**
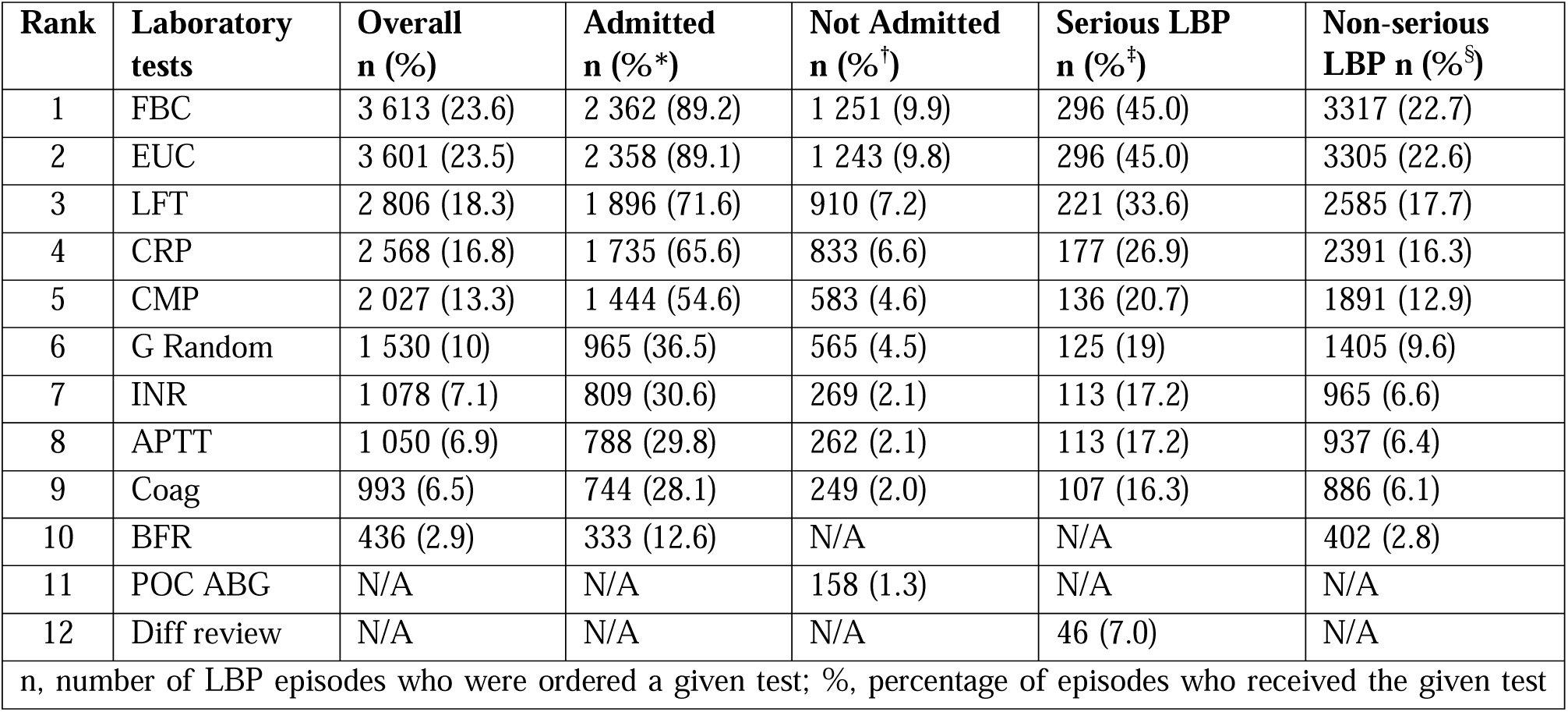
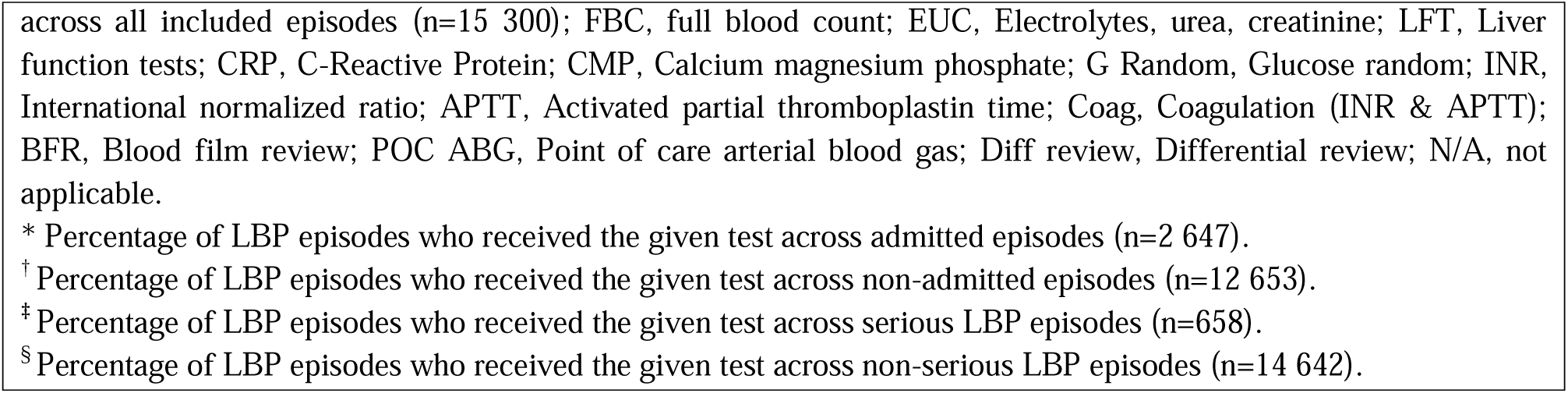
The 10 most commonly ordered laboratory tests.

| Rank | Laboratory tests | Overall n (%) | Admitted n (%*) | Not Admitted n (%†) | Serious LBP n (%‡) | Non-serious LBP n (%§) |
| --- | --- | --- | --- | --- | --- | --- |
| 1 | FBC | 3 613 (23.6) | 2 362 (89.2) | 1 251 (9.9) | 296 (45.0) | 3317 (22.7) |
| 2 | EUC | 3 601 (23.5) | 2 358 (89.1) | 1 243 (9.8) | 296 (45.0) | 3305 (22.6) |
| 3 | LFT | 2 806 (18.3) | 1 896 (71.6) | 910 (7.2) | 221 (33.6) | 2585 (17.7) |
| 4 | CRP | 2 568 (16.8) | 1 735 (65.6) | 833 (6.6) | 177 (26.9) | 2391 (16.3) |
| 5 | CMP | 2 027 (13.3) | 1 444 (54.6) | 583 (4.6) | 136 (20.7) | 1891 (12.9) |
| 6 | G Random | 1 530 (10) | 965 (36.5) | 565 (4.5) | 125 (19) | 1405 (9.6) |
| 7 | INR | 1 078 (7.1) | 809 (30.6) | 269 (2.1) | 113 (17.2) | 965 (6.6) |
| 8 | APTT | 1 050 (6.9) | 788 (29.8) | 262 (2.1) | 113 (17.2) | 937 (6.4) |
| 9 | Coag | 993 (6.5) | 744 (28.1) | 249 (2.0) | 107 (16.3) | 886 (6.1) |
| 10 | BFR | 436 (2.9) | 333 (12.6) | N/A | N/A | 402 (2.8) |
| 11 | POC ABG | N/A | N/A | 158 (1.3) | N/A | N/A |
| 12 | Diff review | N/A | N/A | N/A | 46 (7.0) | N/A |
| n, number of LBP episodes who were ordered a given test; %, percentage of episodes who received the given test |  |  |  |  |  |  |

across all included episodes (n=15 300); FBC, full blood count; EUC, Electrolytes, urea, creatinine; LFT, Liver function tests; CRP, C-Reactive Protein; CMP, Calcium magnesium phosphate; G Random, Glucose random; INR, International normalized ratio; APTT, Activated partial thromboplastin time; Coag, Coagulation (INR & APTT); BFR, Blood film review; POC ABG, Point of care arterial blood gas; Diff review, Differential review; N/A, not applicable.
\* Percentage of LBP episodes who received the given test across admitted episodes (n=2 647).
† Percentage of LBP episodes who received the given test across non-admitted episodes (n=12 653).
‡ Percentage of LBP episodes who received the given test across serious LBP episodes (n=658).
§ Percentage of LBP episodes who received the given test across non-serious LBP episodes (n=14 642).

## Discussion

Nearly a quarter of LBP episodes in participating EDs resulted in at least one pathology test being ordered. We found differences in demographic and clinical characteristics between LBP episodes where pathology testing was ordered compared to where it was not. After adjusting for age, sex and triage category, the factors that had an important influence on pathology test ordering were admission to hospital, a prolonged ED length of stay, arrival to ED by ambulance and a serious LBP diagnosis at discharge from ED. Biochemistry, haematology and immunology were the most frequently ordered categories of laboratory tests, and full blood count, electrolytes, urea, creatinine and liver function tests were the most frequently ordered individual tests. Patients diagnosed with serious LBP or admitted to hospital received higher numbers of pathology tests than patients without these clinical features. A strength of this study is that it included real-world data of a large sample of patients over a 6-year period. To our knowledge, it is the first study to comprehensively describe the profile of adult patients with LBP for whom pathology tests are ordered in EDs, as well as the types of tests ordered.

The strong association we found between admission and pathology testing for patients with LBP is guideline-adherent (5) and consistent with studies in both public and private Australian EDs (12, 13). As in our study, de Luca et al found ambulance arrival to increase the odds of pathology tests being ordered to patients with LBP in EDs (12). In their study on ED care of patients with LBP, Ferreira et al found ambulance arrival and a longer stay in ED to increase the odds of lumbar imaging being ordered (2), as these factors did for pathology testing in our study. A 7-year US national survey revealed that most critically ill patients use ambulance as a transport to ED (20), which could explain ambulance to be a driver for diagnostic work-up. The increased odds of a pathology test being ordered with a prolonged stay in ED is concordant with a 3-year US data set of ED care in which any testing was shown to increase the length of stay, with blood tests being in the top most time consuming tests (21). Serious LBP diagnosis at discharge from ED increased the odds of a pathology test being ordered in our study, which is indirectly guideline-adherent, as pathology test ordering should be guided by the provisional diagnosis at arrival to ED (6). This suggests that pathology testing was used to diagnose serious pathologies.

Our overall 23.8% rate of pathology test ordering closely resembles Nunn et al’s (22.5%) (11). However, they solely examined the tests ordered to patients with LBP in the ED, whereas we included pathology tests ordered in the ED and after inpatient admission. Most pathology tests were ordered for admitted patients in our study, suggesting that fewer pathology tests were ordered in the ED compared to Nunn et al’s study (11). Full blood count and electrolytes, urea, creatinine were each ordered for 24% of our included LBP episodes, which is higher than the rates obtained by Friedman et al in their LBP ED care study (9.7% and 5.2%, respectively) (10). Again, this could be explained by the fact that we included pathology test orders from the ED and the inpatient wards. A notable difference in our findings is that urinalysis was not in the top ten of the most ordered tests, indicating that it was ordered for less than 3% of LBP episodes, whereas it was ordered for 21.9% of patients in Nunn et al (11), and for 18.8% of patients in Friedman et al (10), and it was recommended in guidelines for pathology testing (5, 6). A potential explanation of this finding is that urine dipstick might have been performed by nursing staff as a screening test at arrival to ED, and this information was only recorded in our data set if the result to the test was positive and a formal urinalysis was performed.

Full blood count and electrolytes, urea, creatinine were both ordered for 89%, random glucose for 37%, and urinalysis for less than 13% of admitted episodes. This follows partly the Australian pathology testing guideline by the Colleges for Emergency Medicine and Pathology as all these tests are recommended for admitted patients with non-traumatic LBP (5). INR, APTT, and coagulation studies were ordered for ∼30% of admitted episodes, and liver function tests for 72%. This diverges from the guideline as these tests are generally not indicated (5). All these tests were ordered for non-admitted episodes as well, but with order rates below 10%. We think this could be guideline-concordant as non-admitted episodes were considerably less likely to receive a pathology test than admitted episodes (∼90%), and we did not have the information on pathology suspicion at arrival to ED. LBP episodes who had a diagnosis of a serious pathology at discharge from ED were ordered full blood count, INR, APTT and coagulation studies, electrolytes, urea, creatinine, and liver function tests at higher rates than those who had a non-serious diagnosis.

This suggests concordance with the Australian Emergency Care Institute guideline as these tests are recommended when a non-spinal cause or serious pathology is suspected, but we cannot confirm this hypothesis as we only had the final diagnosis (6). LBP episodes who were admitted or had a serious LBP received a higher number of distinct pathology tests than their counterparts, which is consistent with guidelines (3, 5, 6).

### Limitations

A limitation of the data set is that it only included the final discharge diagnosis and not the competing provisional diagnoses or comorbidities a clinician may have been considering when ordering the tests. We could not determine whether the tests for admitted episodes were ordered in the ED or after inpatient admission, as the data were pooled. These limitations prevent assessing the appropriateness of the ordered tests. Given the study’s retrospective nature, misclassification bias might have occurred for some variables (e.g. admission status). There is uncertainty regarding the generalisability of our findings to other settings such as primary care, where serious LBP pathologies have lower prevalences than in the ED (22), or to other countries. Another limitation is that we did not have the information on the tests that were performed during the screening by the nursing staff unless they were sent to the laboratory. For example, dipstick urinalysis was only sent to the pathology department for analysis when it was positive for nitrites and/or leucocytes (23).

### Conclusions

We could not make a definitive judgement on the appropriateness of the pathology test ordering in our study. Yet, we believe that increased odds of pathology test orders when patients were admitted, had a serious diagnosis of LBP, and were transported by ambulance are guideline-adherent (5, 6). We might question the high ordering rates of liver function tests and coagulation studies and the very low rates of urinalysis for admitted patients, as these do not follow the recommendations (5). However, we did not have the information on dipstick urinalysis that was performed as a screening test by the nursing staff, so we cannot confirm that the low rate of urinalysis reflects non-adherence to guidelines. Future studies should examine the appropriateness of pathology testing for patients with LBP in EDs and other settings to screen for low-value care, potentially decrease the length of stay and improve LBP management.

## Acknowledgements

CCP is supported by a scholarship from the Quebec Research funds – Health division. GCM and CGM are supported by Investigator Grants from the Australian National Health and Medical Research Council (NHMRC). HMA is supported by a Junior 2 research scholar from the Quebec Research Funds – Health division.

## Statements and Declarations

### Data availability statement

Data are available on reasonable request.

### Author contributions

Study conception and design: CCP, QC, HMA, PY, MS, CGM, GCM; Data curation: QC, GCM; Formal analysis: CCP, QC, HMA, GCM; Project administration: CCP, GC, CGM; Writing – original draft: CCP; Writing – review and editing: CCP, QC, HMA, PY, MS, CGM, GCM.

## References

1. Edwards J, Hayden J, Asbridge M, Gregoire B, Magee K. Prevalence of low back pain in emergency settings: a systematic review and meta-analysis. BMC Musculoskelet Disord. 2017;18(1):143. Epub 2017/04/06. doi: 10.1186/s12891-017-1511-7. PubMed PMID: 28376873; PubMed Central PMCID: PMCPMC5379602.

2. Ferreira GE, Machado GC, Abdel Shaheed C, Lin CC, Needs C, Edwards J, et al. Management of low back pain in Australian emergency departments. BMJ Qual Saf. 2019;28(10):826–34. Epub 2019/06/06. doi: 10.1136/bmjqs-2019-009383. PubMed PMID: 31164487.

3. Chou R, Qaseem A, Snow V, Casey D, Cross JT, Jr., Shekelle P, et al. Diagnosis and treatment of low back pain: a joint clinical practice guideline from the American College of Physicians and the American Pain Society. Ann Intern Med. 2007;147(7):478–91. Epub 2007/10/03. doi: 10.7326/0003-4819-147-7-200710020-00006. PubMed PMID: 17909209.

4. Oliveira CB, Maher CG, Pinto RZ, Traeger AC, Lin CC, Chenot JF, et al. Clinical practice guidelines for the management of non-specific low back pain in primary care: an updated overview. Eur Spine J. 2018;27(11):2791–803. Epub 2018/07/05. doi: 10.1007/s00586-018-5673-2. PubMed PMID: 29971708.

5. ACEM, RCPA. Pathology testing in the emergency department developed by the Australasian College for Emergency Medicine (ACEM) and the Royal College of Pathologists of Australasia (RCPA) 2023 [cited 2024 June 4]. Available from: https://acem.org.au/getmedia/57501811-e932-4c74-85be-159f0621917f/RCPA-ACEM-Guideline-v01-(Mar-13)-Final.aspx.

6. ECI. Emergency Care Institute NSW - Acute low back pain aci.health.nsw.gov.au [cited 2024 June 4]. Available from: https://aci.health.nsw.gov.au/networks/eci/clinical/clinical-tools/orthopaedic-and-musculoskeletal/acute-low-back-pain#i.

7. Downie A, Hancock M, Jenkins H, Buchbinder R, Harris I, Underwood M, et al. How common is imaging for low back pain in primary and emergency care? Systematic review and meta-analysis of over 4 million imaging requests across 21 years. Br J Sports Med. 2020;54(11):642-51. Epub 2019/02/15. doi: 10.1136/bjsports-2018-100087. PubMed PMID: 30760458.

8. Traeger AC, Machado GC, Bath S, Tran M, Roper L, Oliveira C, et al. Appropriateness of imaging decisions for low back pain presenting to the emergency department: a retrospective chart review study. Int J Qual Health Care. 2021;33(3). Epub 2021/07/15. doi: 10.1093/intqhc/mzab103. PubMed PMID: 34260690.

9. Mau AWK, Keen HI, Hill CL, Buchbinder R. Appropriateness of lumbar spine imaging in patients presenting to the emergency department with low back pain in a Western Australian tertiary hospital. Intern Med J. 2025. Epub 2025/01/04 20:12. doi: 10.1111/imj.16626. PubMed PMID: 39754748.

10. Friedman BW, Chilstrom M, Bijur PE, Gallagher EJ. Diagnostic testing and treatment of low back pain in United States emergency departments: a national perspective. Spine (Phila Pa 1976). 2010;35(24):E1406-11. Epub 2010/10/30. doi: 10.1097/BRS.0b013e3181d952a5. PubMed PMID: 21030902; PubMed Central PMCID: PMCPMC2982879.

11. Nunn ML, Hayden JA, Magee K. Current management practices for patients presenting with low back pain to a large emergency department in Canada. BMC Musculoskelet Disord. 2017;18(1):92. Epub 2017/02/24. doi: 10.1186/s12891-017-1452-1. PubMed PMID: 28228138; PubMed Central PMCID: PMCPMC5322663.

12. de Luca K, McLachlan AJ, Maher CG, Machado GC. Australian emergency department care for older adults diagnosed with low back pain of lumbar spine origin: a retrospective analysis of electronic medical record system data (2016-2019). BMC Emerg Med. 2023;23(1):17. Epub 2023/02/15. doi: 10.1186/s12873-023-00789-8. PubMed PMID: 36782123; PubMed Central PMCID: PMCPMC9924838.

13. Buchbinder R, Bourne A, Staples M, Lui C, Walker K, Ben-Meir M, et al. Management of patients presenting with low back pain to a private hospital emergency department in Melbourne, Australia. Emerg Med Australas. 2022;34(2):157–63. Epub 2021/06/25. doi: 10.1111/1742-6723.13814. PubMed PMID: 34164911.

14. Hansen DP, Kemp ML, Mills SR, Mercer MA, Frosdick PA, Lawley MJ. Developing a national emergency department data reference set based on SNOMED CT. Med J Aust. 2011;194(4):S8–10. Epub 2011/03/16. doi: 10.5694/j.1326-5377.2011.tb02934.x. PubMed PMID: 21401491.

15. Benchimol EI, Smeeth L, Guttmann A, Harron K, Moher D, Petersen I, et al. The REporting of studies Conducted using Observational Routinely-collected health Data (RECORD) statement. PLoS Med. 2015;12(10):e1001885. Epub 2015/10/07. doi: 10.1371/journal.pmed.1001885. PubMed PMID: 26440803; PubMed Central PMCID: PMCPMC4595218 conflicts of interest to declare.

16. Machado GC, O’Keeffe M, Richards B, Needs C, Storey H, Maher CG. Why a dearth of sports and exercise medicine/physiotherapy research using hospital electronic medical records? A success story and template for researchers. Br J Sports Med. 2020. Epub 2020/05/23. doi: 10.1136/bjsports-2019-101622. PubMed PMID: 32439629.

17. ABS. Australian Bureau of Statistics’ Socio-Economic Indexes for Areas (SEIFA) 2016 [cited 2024 May]. Available from: https://www.abs.gov.au/ausstats/abs@.nsf/mf/2033.0.55.001.

18. Ebrahimi M, Heydari A, Mazlom R, Mirhaghi A. The reliability of the Australasian Triage Scale: a meta-analysis. World J Emerg Med. 2015;6(2):94–9. Epub 2015/06/10. doi: 10.5847/wjem.j.1920-8642.2015.02.002. PubMed PMID: 26056538; PubMed Central PMCID: PMCPMC4458479.

19. Sullivan GM, Feinn R. Using Effect Size-or Why the P Value Is Not Enough. J Grad Med Educ. 2012;4(3):279–82. Epub 2013/09/03. doi: 10.4300/JGME-D-12-00156.1. PubMed PMID: 23997866; PubMed Central PMCID: PMCPMC3444174.

20. Squire BT, Tamayo A, Tamayo-Sarver JH. At-risk populations and the critically ill rely disproportionately on ambulance transport to emergency departments. Ann Emerg Med. 2010;56(4):341–7. Epub 2010/06/18. doi: 10.1016/j.annemergmed.2010.04.014. PubMed PMID: 20554351.

21. Kocher KE, Meurer WJ, Desmond JS, Nallamothu BK. Effect of testing and treatment on emergency department length of stay using a national database. Acad Emerg Med. 2012;19(5):525–34. Epub 2012/05/19. doi: 10.1111/j.1553-2712.2012.01353.x. PubMed PMID: 22594356.

22. Galliker G, Scherer DE, Trippolini MA, Rasmussen-Barr E, LoMartire R, Wertli MM. Low Back Pain in the Emergency Department: Prevalence of Serious Spinal Pathologies and Diagnostic Accuracy of Red Flags. Am J Med. 2020;133(1):60–72 e14. Epub 2019/07/07. doi: 10.1016/j.amjmed.2019.06.005. PubMed PMID: 31278933.

23. ECI. Emergency Care Institute, Acute lower back pain - Adult ECAT protocol 2023 [cited 2025 March 20]. Available from: https://aci.health.nsw.gov.au/ecat/adult/acute-lower-back-pain.

