## Supplementary Table 1 for "Pathology testing for patients with low back pain in Australian emergency departments"

**Supplementary Table 1.** Label of tests that were included in each category and of excluded variables.

| **Categories** | **Name of tests** |
| --- | --- |
| 1. Biochemistry | _Ur Creatinine Shared, _Urine Creatinine Analysis Random, _Urine Microalbumin/Creat Ratio Random, Active B12 (Holotranscobalamin) Level, Albumin, Alkaline Phosphatase, Alpha 1 Antitrypsin, Ammonia, Amylase, Angiotensin Converting Enzyme, Aspartate Aminotransferase, Bilirubin, Calcium, Calcium Magnesium Phosphate, Ceruloplasmin, Cholesterol, Creatine Kinase, Creatine Kinase Electrophoresis, CSF Glucose, CSF Protein and Glucose, Electrolytes Glucose Creatinine - SLHD only, Electrolytes Liver Function, Full Blood Count, Electrolytes Urea Creatinine, EUC and LFT, G6PD Screen, Glucose Fasting, Glucose Random, High Density Lipoprotein Cholesterol, Iron, Iron Studies, Lactate Dehydrogenase, Lipase Analysis, Lipids, Liver Function Tests, Low Density Lipoprotein Cholesterol, Magnesium, Osmolality, Phosphate, Potassium, Ser Folate, Serum Folate, Triglycerides, Troponin T - high sensitive, Urate, Urea, Urine Creatinine Random, Urine Microalbumin/Creat Ratio Random, Urine Osmolality Random, Urine Protein Random, Urine Sodium and Osmolality Random, Urine Sodium Random, Vit B12 and Serum Folate, Vit B12 Serum Folate and Iron Studies, Vitamin B12 and Serum Folate, Vitamin B12 Level, Vitamin B12, Serum Folate and Iron Studies, Vitamin D 125 Dihydroxy Level, Vitamin D 25 Hydroxy Level. |
| 2. Haematology | _bloodfilmfiled, _bloodfilmreview, _bloodfilmreviewtracking, _bloodfilmreviewtrackingaliquot, _Differential Review Tracking Aliquot, _differentialreview, _differentialreviewtracking, _Haemoglobin A2, _Manual Differential Statistics, Beta 2 Microglobulin, Bone Marrow Chromosome Studies, Bone Marrow Molecular Cytogenetics FISH, Bone Marrow Surface Markers, Cancellation Haematology, Erythrocyte Sedimentation Rate, Full Blood Count, Haematology Lymphoma Screen – CRGH, Haematology Myeloma Screen – CRGH, Haematology Profile, Haemoglobin F, Haptoglobin, Lymphocytes Subsets, Reticulocytes, Thalassaemia Screen (HbA2,HbF). |
| 3. Immunology | _EPG Total Protein – EZ, _EPG Total Protein Check – EZ, _Immunoelectrophoresis. _Protein Electrophoresis, _Serum Protein Electrophoresis Check, _Urine Protein Electrophoresis Random, Acetylcholine Receptor Antibody, Anti GQ1B Ab, Antinuclear Antibodies, C Reactive Protein, Complement C2, Complements C3 and C4, Cyclic Citrullinated Peptide Antibodies, Double Stranded DNA Antibodies Assay, Endomysial Antibodies IgA, Extractable Nuclear Antigens Ab, Glomerular Basement Membrane Antibodies, Human Leucocyte Antigen B27, Immunoelectrophoresis, Immunofixation Electrophoresis, Immunoglobulin A Level, Immunoglobulin G Level,  Immunoglobulin M Level, Immunoglobulins A G and M Level, Liver Kidney Microsomal Antibodies, Muscle Specific Kinase Antibody, Neutrophil Cytoplasmic Antibodies, Protein Electrophoresis – EZ, Rheumatoid Factor, Serum Free Light Chains, Smooth Muscle Antibodies, Transglutaminase IgA Antibodies, Urine Protein Electrophoresis Random - EZ. |
| 4. Coagulation | APTT, Cancellation Coagulation, Clexane anti-Xa activity, Coagulation (INR & APTT), D-Dimer Assay, Factor X Level, Fibrinogen, Fragmin Level anti-Xa activity, INR, Thrombin Time. |
| 5. Blood Gas | Arterial Blood Gas, Blood Gas Venous, POC ABG, Venous Blood Gas, Venous Blood Gas Lactate, Venous Blood Gas plus Lactate, Venous Blood Gas Potassium, Venous Blood Gases. |
| 6. Endocrinology | Adrenocorticotropic Hormone Level, Aldosterone, Beta HCG Pregnancy Assay, Cortisol Level, CSF Beta HCG, Free Thyroxine (Abbott), Glycosylated Haemoglobin A1c ?  Diabetic, Glycosylated Haemoglobin A1c known Diabetic, Parathyroid Hormone, Progesterone Level, Prolactin, Prostate Specific Antigen Free, Prostate Specific Antigen Total, Renin Activity, Testosterone Level, Thyroid Function Tests, Thyroid Microsomal Antibodies, Thyroid Stimulating Hormone, Thyroxine Free, Triiodothyronine Free, Urine Beta HCG Pregnancy Screen. |
| 7. Toxicology | Apixaban Level, Carbamazepine Level, Digoxin Level, Ethanol Level, Everolimus Level, Lamotrigine Level, Levetiracetam Level, Lithium Level, Paracetamol Level, Phenytoin Level, Salicylate Level, Tacrolimus Level, Urine Drug Screen Immunoassay, Urine Drug Screen Testing (CRGH only), Valproate Level. |
| 8. Microbiology | _chlamydiatrachomatisnucleicacid, _N.gonorrhoeae Nucleic Acid Detection, _Respiratory Nucleic Acid Detection, Brucella Serology, Chlamydia and N.gonorrhoeae DNA Detection, COVID-19 Diagnostic Orders, CSF Cryptococcal Antigens, CSF Meningitis Nucleic Acid Detection, Cytomegalovirus Acute Infection Serology, Cytomegalovirus Immune Status Serology, Dengue virus Serology, Enterovirus RNA Detection, Epstein Barr virus IgM Antibodies, Epstein Barr virus Serology, Epstein-Barr virus Abs Acute Infection, Faeces Bacterial DNA Screen, Faeces Bacterial Screen, Hepatitis A Antibody Total, Hepatitis B Core Antibodies Total, Hepatitis B DNA Assay,  Hepatitis B Surface Antibodies, Hepatitis B Surface Antigens, Hepatitis C Antibodies, Hepatitis C Genotyping, Hepatitis C RNA Assay NAD, Hepatitis C RNA Screen, Hepatitis C Supplementary Enzyme Immunoassay, Herpes simplex virus Abs Acute Infection, Herpes simplex virus DNA Detection, HIV 1 and 2 Antibodies Status, HIV 1 Viral Load, Infectious Mononucleosis Screen, Parvovirus Serology, Respiratory Pathogens Nucleic Acid Detection, Respiratory Rapid Nucleic Acid Detection, Ross River Virus Serology, SARS-CoV-2 Nucleic Acid Detection, SARS-CoV-2 Nucleic Acid Supplementary, SARS-CoV-2 Rapid Nucleic Acid Detection, Strongyloides Serology, Syphilis Antibodies Screen EIA, TB Interferon Gamma Release Assay, Urine Casts Red Cell Morphology, Urine Microscopy, Varicella zoster Abs Acute Infection, Varicella zoster DNA Detection, Varicella zoster Serology Immune Status, WZ Urine Microscopy. |
| 9. Oncology | Alphafetoprotein Tumour Markers, Beta HCG Tumour Markers, Cancer Associated 125 Antigens, Cancer Associated 15.3 Antigens, Cancer Associated 19.9 Antigens, Carcinoembryonic Antigen. |
| Excluded variables | _Fasting Status, _POC CRG Billing, _POC Liat, _POC RPA Billing, _Slide, Add on Lab Test(s), Chemistry Comment, Coagulation Comment, DIC Screen, ED # NOF/Elderly #, ED Abdo Pain/Liver Diseases/GIT Bleeding – CANT, ED Abdominal Pain, ED Back Pain, ED Cardiac, ED Confusion, ED COVID-19 Initial Orders, ED General, ED General – Cant, ED Geriatric, ED Liver Disease/GIT Bleeding, ED Overdose, ED Profile, ED PV Bleeding, ED Rheumatology, ED Seizures, ED Sepsis, ED SOB, ED TIA/Stroke, ED Trauma, Haem Comment, ICU Routine Profile, Notify, Rheumatology Routine Profile, Spare EDTA, Spare Gel, Spare LiGreen, Spare Red, Spare Tube. |
